# Comparison of epidemic control strategies using agent-based simulations

**DOI:** 10.1101/2020.05.24.20111625

**Authors:** Pablo Martínez Ruiz del Árbol, Lara Lloret Iglesias

**Affiliations:** Instituto de Física de Cantabria

## Abstract

A simulation of the dynamics of a small population is used to assess the impact of different confinement and testing strategies in the control of an epidemic. The simulation considers individuals as agents moving randomly across the habitat according to predefined urban patterns. Agents carry a simple tracing device that identifies signals emitted by other agents, recording the position and time of the encounter. The information of every device is propagated daily to an epidemic observatory based on an online graph database. Infections are simulated as stochastic processes depending on the proximity among individuals. Different epidemic control strategies are tested with and without the information of the tracing device under several scenarios. We observe that the success of the strategies strongly depends on the duration of the period of infectiousness before the presence of symptoms and the fraction of asymptomatic agents. If these values are high, strategies based on the presence of symptoms or on testing campaigns can hardly contain the epidemic. Strategies using massive confinement of the agents are able to control the epidemic at the cost of sending a large fraction of the population into quarantine. In cases with moderate and low values for these parameters, the tracing devices can provide a slightly better performance but only if a large fraction of the agents carry the device. Otherwise, the impact of these devices is found to be negligible in comparison with other strategies not using them. Finally, we provide a methodology allowing to use the information of the graph database to estimate basic parameters of the disease such as the infection probability.

## 1 Introduction

The COVID-19 outbreak was first identified in Wuhan (China) in December 2019 [1]. On the 30th January 2020, the outbreak was declared to be a Public Health Emergency of International Concern by the WHO [2] and a pandemic on the 11th of march 2020 [3]. Since then, apart from the impact in the population health and the numerous casualties, the COVID-19 epidemic has generated an unprecedented shock to the global economy through the severe social distancing and lock-down restrictions imposed by the governments. In such a scenario, the debate was triggered on the effectiveness of the contingency measures and how they could be improved, becoming obvious that there is a need to understand which are the most useful tools to control the disease spread. The ability of a country to handle such a health emergency relies on many different factors: the decisive actions taken by the governments, the population compliance to the restrictions and the capacity to perform adequate and meaningful testing campaigns to the population. Related with the latter, there has been great controversy regarding the utility of contact tracing using smartphone applications. If two smartphones with the app installed are in the proximity, they exchange information and create a contact log. When one of the app users is detected as infected, either because of presenting clear symptoms or because of a positive test, the contact log can be used to quickly identify potentially infected people and apply some action on them such as performing tests or recommending a preventive isolation. Many countries are developing such apps but, leaving privacy considerations aside [4], the debate on their utility is still on the table. South Korea and Singapore are the most quoted examples of the usage of such applications, but their real impact is not clear and the available information is contradictory and incomplete [5, 6]. What seems to be clear is that one of the main caveats of the contact tracing is related with the voluntary use of app. For this approach to be effective, it is obvious that the percentage of population engagement is one of the key parameters. Actually, the greatest engagement among the countries already using this kind of apps is only of about 20%, while the experts claim that at least a 60% engagement is needed to be useful. According to 2019 data only 76% of the people in Europe have mobile internet subscriptions[7], being this number even lower among the elderly people, who happen to be the most vulnerable to the COVID-19 disease. Other parameters affecting the effect of the contingency strategies are more related with the particular characteristics of the disease itself, such as the infectious time before the symptoms appear, the number of asymptomatic and the infection probabilities, among others. Since we are still immersed in the COVID-19 emergency, many of these parameters are not yet well known. The goal of this work is to present several scenarios with different possible disease parameters and study the impact of six epidemic control strategies (ECS), some of them including contact tracing information.

## 2 Epidemic modeling using agent-based simulations

The epidemic dynamics have been modelled using autonomous agents interacting with each other in the context of a closed environment referred to as *habitat*. The simulation considers the time as a discrete variable with a granularity of 1 minute.

### Habitat modeling

The population is distributed over a squared grid of blocks of equal sizes. The blocks can have three differential purposes: residential buildings (RB), work places (WP) or places dedicated to social activities (social places, SP). The fraction of each type is a configurable parameter of the habitat. Both RB and WP have several floors according to a Poisson distribution, while the SP have one floor in all cases. Each of the floors in a block is divided in several apartments according to a normal distribution. The number of inhabitants per apartment is modeled as a Poisson distribution. All the apartments have a set of locations with a higher probability of being used by the inhabitants. These locations are known as frequent points (FP). The number of FPs is obtained from a Poisson distribution and their position from a uniform distribution limited to the extension of the apartment. An example of this spatial arrangement is shown in Fig. 1.

**Figure 1:**
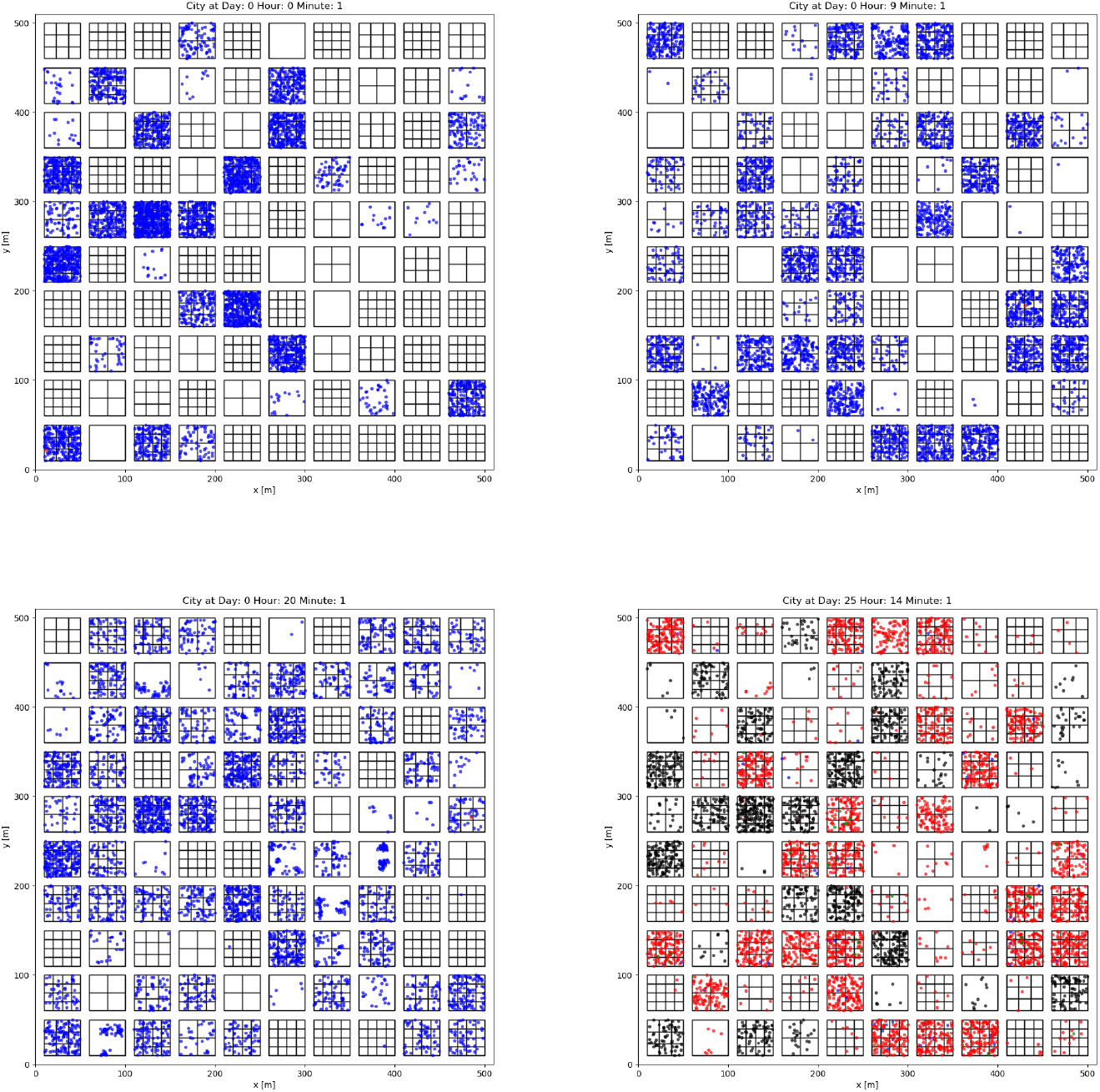
Floor plan of a population habitat at different times of the epidemic progression. The buildings are represented as boxes containing several floors (not seen in this projection) with apartments represented as sub-boxes. Agents are represented as dots with colors indicating their medical status: blue if susceptible, red if infected, green if cured and black if in quarantine. The upper and lower-left figures illustrate day one at three different times in which the agents are mainly at the RBs, WPs and SP respectively. The lower-right figure shows a case in which the epidemic has progressed for many days.

### Agent modeling

The logic of the agents consists of three components: the city location scheduler, the health state manager, and the contact tracing system (CTS). Agents are first randomly assigned to the RBs according to the habitat structure. Each of the agents is also assigned a unique workplace and a set of preferred SPs. The location scheduler considers a day-cycle with three phases: pernoctation at the RBs, work activity at the WPs, and social activity at the SPs. The times at which there is a transition between phases are extracted from normal distributions, changing for every agent and day. Agents can freely move inside the apartments. The time that every agent stays at a given position at the apartment is taken randomly from a Poisson distribution, different for RBs, WPs and SPs. When this time expires a new position within the apartment is assigned first by randomly selecting one of the FPs, and then according to a normal distribution around it. Agents in the social activity phase can also commute among their preferred SPs, although a small probability exists to commute to any other SP in the habitat. The length of the staying at each SP is taken from a Poisson distribution.

Agents have three different medical states: susceptible, infected or recovered. At the beginning of the epidemic one agent is initialized as infected while the rest are initialized as susceptible. The behavior of the disease in each agent is determined by several parameters chosen randomly. An agent can be symptomatic or asymptomatic accordingly to a configurable parameter expressing the expected fraction of asymptomatic cases. Infections are modeled as random events, with a user-defined constant probability, any time that two agents are closer than a configurable distance during one minute. The time since an agent is infected and it can infect other agents, the time since the agent can infect and the symptoms appear (if symptomatic), and the time until the agent is considered recovered and can not propagate the disease, are taken randomly for each agent from Poisson distributions. Agents with symptoms or under prescription according to some epidemic-control strategy (ECS) can be declared as being in quarantine. If this state is activated the agent can not infect other agents and the day-cycle of the agent is restricted to being at its corresponding RB, until the state is revoked.

All agents are programmed to carry a simulated CTS. Agents closer than a configurable distance to any other agent establish a proximity contact characterized by the identification code of the other agent and the location, time and duration of the contact. If the proximity contact extends in time, the contact information is updated including the total duration of the event. All this information is then stored in a graph database described in more details in the next section. The fraction of agents actually porting such a CTS is given as a parameter to the simulation.

### Epidemic observatory using a graph database

All the information concerning the medical state of the agents (if known) and their contact trace history is given as input to an online graph database. This choice is justified by the nature of the infection process which depends on the proximity contacts between pairs of individuals. Agents using a CTS, have a random, predefined, daily time in which a communication is established with the online database. The two agents involved in the proximity contact are inserted in the database as vertices and the contact itself as an edge connecting the two. The vertices store information on the current medical status of the agents while each of the edges contains the time, location and duration of the contact. This study has been performed using a *JanusGraph* [8] database, since it is optimized for storing and querying graphs containing billions of vertices and edges distributed across machine clusters, supporting thousands of concurrent users executing complex graph traversals in real time. Additionally, *JanusGraph* is massively scalable and could be transparently adapted to be used on larger populations. A visual representation of a the graph database for a small population of about 369 inhabitants after 12 days of simulation can be seen in Fig. 2. The database allows to easily select an infected agent and to automatically get the list of the last agents involved in an interaction with it. This list can also be classified in terms of the frequency and duration of the contacts. The quarantine state of the agents can be modified daily by an epidemic-control logic which implements different strategies with and without the information of the CTS. These strategies are described in the next section. The data analysed in this work have been extracted from the database information filled during the simulations.

**Figure 2:**
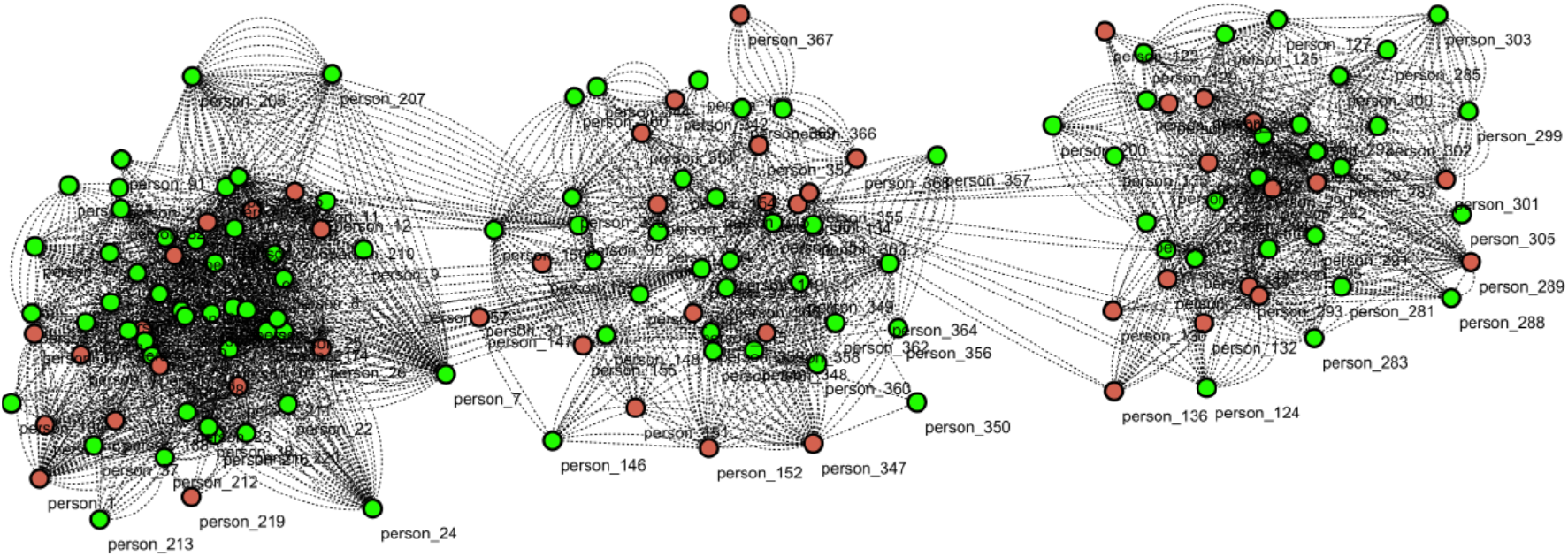
Visual representation of the graph generated in a simulation with 369 agents. The people marked in green are healthy (or not yet detected as infected) agents and the red nodes correspond to the confirmed infected agents.

## 3 Implementation of epidemic control strategies

Six ECS have been tested using different criteria to send people into quarantine. Its logic is implemented using the information of the graph database: the presence or absence of symptoms, the results of a testing campaign, and the CTS logs. Strategies using CTS are especially interesting given the lack of knowledge about their effectiveness, although approaches not using this information are also studied to clearly establish a set of benchmarks aiming to achieve a fair comparison. The traced contacts for a strategy using CTS refers to the list of agents who entered into the CTS range of an infected agent during the 5 days previous to be tagged as such. Since the number of tests is limited, when the traced contacts are used for testing purposes, they are sorted by the total interaction duration aiming at maximizing the chances of detecting new infected people. The person can be tagged as infected both because of the manifestation of symptoms or because of a positive test. Several scenarios have been studied for the different ECS depending on the number of tests available and the engagement to the tracing application.

The first ECS (ECS0) is the baseline for all the others. This ECS does not consider testing on the population nor the use of any CTS. In this case, agents go into quarantine only when (if) symptoms manifest. The second ECS (ECS1) extends ECS0 by performing random tests on the population with a maximum capacity per day. Agents are set on quarantine if symptoms appear or if they are tested positive. The next ECS (ECS2) is similar to ECS1 but using a more selective testing that targets the cohabitants of infected people who manifested symptoms. The forth ECS (ECS3) considers the use of a CTS to extend the testing to all the traced contacts of an agent that manifested symptoms. The next ECS (ECS4) does not perform testing on the population and simply sends into quarantine when an agent has symptoms or if it belongs to the traced contacts of any other infected agent. The last ECS (ECS5) follows ECS4 but those agents being sent into quarantine are tested and released if the result is negative. In all cases the testing of an agent is not performed if the agent was already tested in the last 5 days. Table 1 presents a summary of these ECS and their characteristics.

**Table 1:**
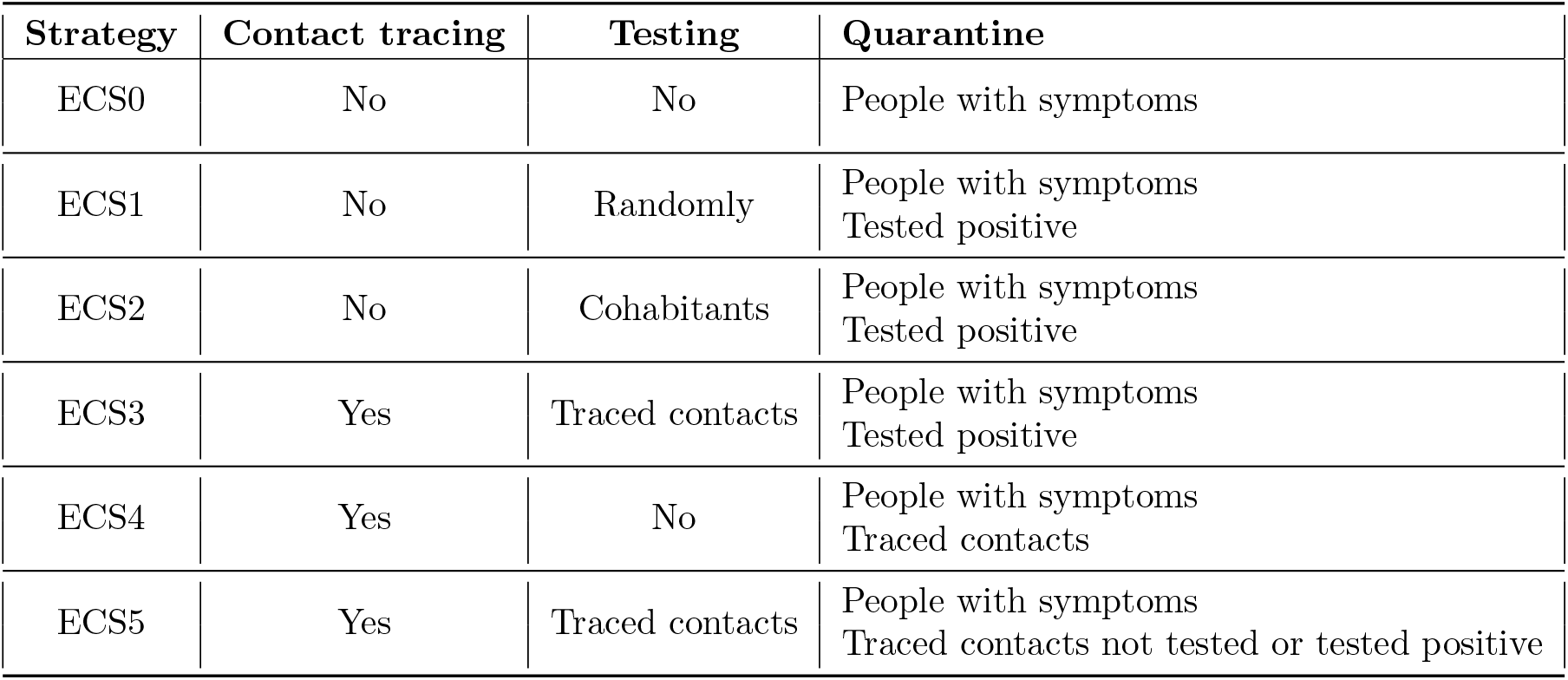
Summary of the strategies proposed in this study according to the confinement regime attempted, and to whether a CTS is used and tests are made on suspicious individuals.

| Strategy | Contact tracing | Testing | Quarantine |
| --- | --- | --- | --- |
| ECS0 | No | No | People with symptoms |
| ECS1 | No | Randomly | People with symptoms<br>Tested positive |
| ECS2 | No | Cohabitants | People with symptoms<br>Tested positive |
| ECS3 | Yes | Traced contacts | People with symptoms<br>Tested positive |
| ECS4 | Yes | No | People with symptoms<br>Traced contacts |
| ECS5 | Yes | Traced contacts | People with symptoms<br>Traced contacts not tested or tested positive |

## 4 Results

### Population and disease parameters setup

This study considers 70-days-long simulations with dynamics driven by the parameters shown in table 2. This configuration corresponds to a population of roughly 10000 inhabitants. In order to account for the fluctuations due to the random dynamics of the epidemic, a total of 10 different seeds have been used for each simulated scenario. This has been proven to be important especially in scenarios with a low infection probability in which the epidemic can be often stopped by the application of the ECS.

**Table 2:** Parameters driving the structure of the population habitat, the mobility of the agents, the disease characteristics and the contact-tracer features. Those parameters with several values have been studied in detail to see their impact on the dynamics of the epidemic.

| Population habitat parameters |  |  |
| --- | --- | --- |
| Parameter | Distribution | Value |
| Surface | Fixed | 36000 $m^2$ |
| Fraction of RB (WP,SP) | Uniform | 0.4 (0.3, 0.3) |
| Number of floors per buildings | Poisson | $\lambda=5$ |
| Number of apartments per floor | Poisson | $\lambda=3$ |
| Number of inhabitants per apartments | Poisson | $\lambda=3$ |
| Number of FPs per apartment | Poisson | $\lambda=4$ |
| Proximity to FPs | Gaussian | $\mu=0, \sigma=3$ m |
| Number of frequent leisure places for agent | Poisson | $\lambda=3$ |
| Mobility parameters |  |  |
| Parameter | Distribution | Value |
| Transition from being at RB and WP | Gaussian | $\mu=9$ h, $\sigma=60$ min |
| Transition from being at WP and SP | Gaussian | $\mu=16$ h, $\sigma=60$ min |
| Transition from being at SP and RB | Gaussian | $\mu=20$ h, $\sigma=120$ min |
| Time scale of changing location inside RB | Poisson | $\lambda=120$ min |
| Time scale of changing location inside WP | Poisson | $\lambda=60$ min |
| Time scale of changing location inside SP | Poisson | $\lambda=30$ min |
| Time scale of changing SP | Poisson | $\lambda=60$ min |
| Disease parameters |  |  |
| Parameter | Distribution | Value |
| Infection probability (30 min) | Fixed | 0.025/0.035/0.045 |
| Infection radius | Fixed | 2 m |
| Latent period | Poisson | $\lambda=2$ days |
| Incubation - Latent period | Poisson | $\lambda=0/2/4$ days |
| Symptomatic period | Poisson | $\lambda=10$ days |
| Fraction of asymptomatic | Uniform | 0.25/0.75 |
| Contact-tracer parameters |  |  |
| Parameter | Distribution | Value |
| Contact-tracer influence radius | Fixed | 2 m |
| Contact-tracer active history logs | Fixed | 5 days |
| Minimum contact duration | Fixed | 10 min |
| Engagement of contact-tracer | Uniform | 0.25/0.75/1 |

The disease-related parameters have been set according to ranges seen in the literature [9, 10]. Three different infection probabilities, expressed as the probability of an agent being infected after being 30 minutes within the infection radius of an infected agent, are considered. These probabilities generate epidemics with *R*_0_ values in the range 2-10. The fraction of asymptomatic agents has been considered to be 25% and 75%. The infectious period before the presence of symptoms (IPS) has been varied between 0, 2 and 4 days. In those strategies involving massive testing campaigns, two testing-capacities of 100 and 300 tests per day have been considered. Finally, the fraction of agents carrying a CTS has been varied between 25%, 75% and 100%. A total of 200 simulations have been produced to perform this study.

### Epidemic evolution and observables

The simulations considered in this work have been run with a granularity of 1 minute during 70 days for every scenario and seed. Six observables have been monitored at every simulation step: the fractions of susceptible, infected and recovered population, the fraction of the population being in quarantine, the cumulative number of tests performed (for ECS using testing), and the final fraction of tests that resulted in a positive outcome. Figure 3 shows an example of these observables for a given scenario and seed.

**Figure 3:**
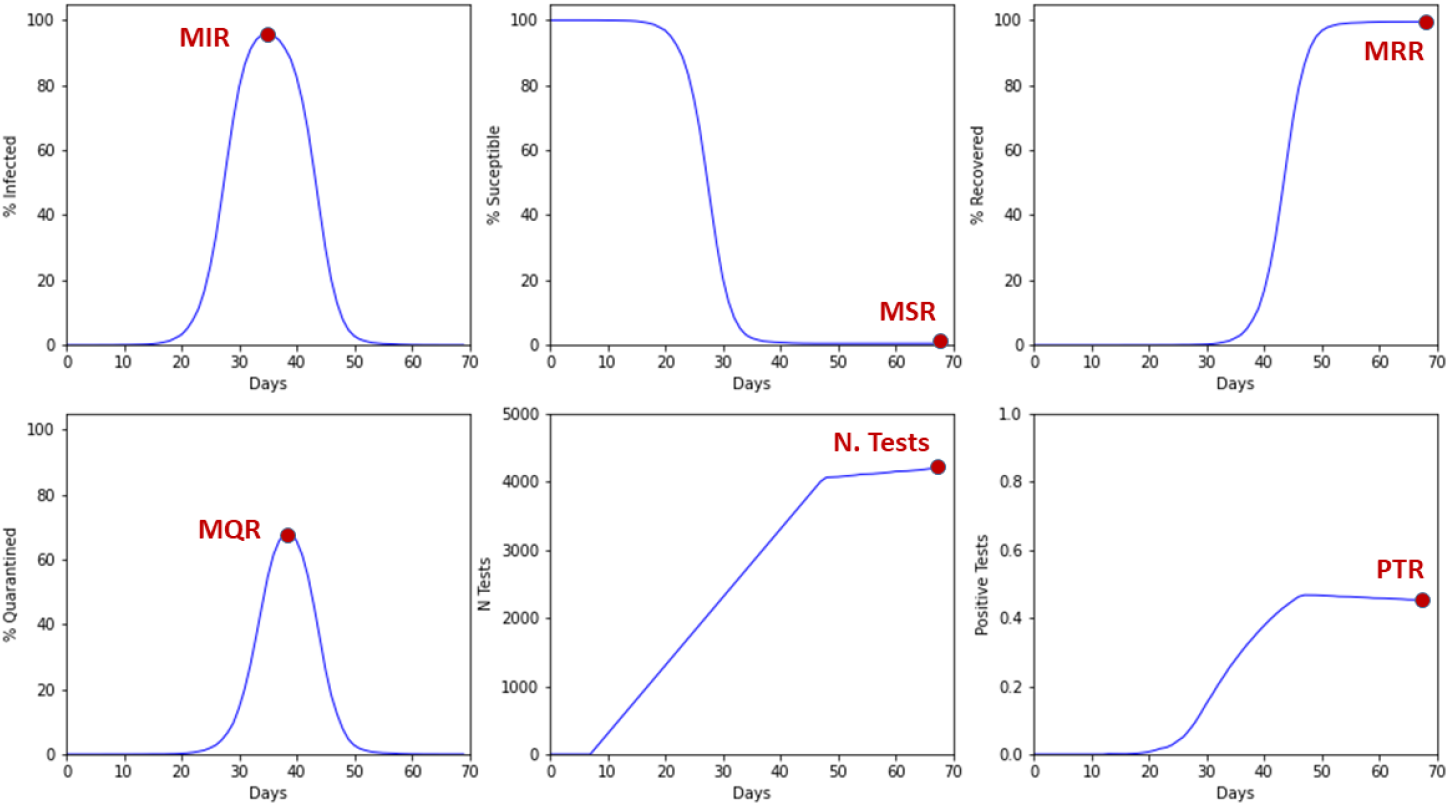
Time evolution of the epidemic for a given scenario and seed. The fraction of infected, susceptible, and recovered agents, the fraction of agents in quarantine, the cumulative number of tests and the fraction of positive tests are shown. The maximum values (minimum in the case of susceptible) is use as the metric to compare among different ECS.

The comparisons among different scenarios are performed using the extreme values of the epidemic evolution. These observables are: the minimum susceptible rate (MSR), the maximum infected rate (MIR), the maximum recovered rate (MRR), the maximum rate of quarantined population (MQR), the total number of tests (N.Tests), and the total rate of positive tests (PTR).

### Dependency on the infection probability

Figure 4 shows the score of the different ECS for increasing infection probabilities. The first conclusion that we can draw after inspecting the plots in the figure, is that we can divide the ECS into two groups according to their behaviour: strategies where quarantine only happens after the presence of symptoms or after positive testing (ECS 0 to 3) and strategies where the quarantine is massively applied to suspicious agents (ECS 4 and 5). The former group yield relatively similar results in terms of the MIR, always ranging between to 80% to 100% for all strategies. This shows that for such a large IPS most of the strategies have a mild impact on the disease spread because by the time an infected agent is detected, it was already able to freely propagate the disease for a few days. It can be observed that, in the low probability case, the epidemic can be stopped on time for some seeds, occurring more often as the strategy number increases. Concerning the MQR we observe similar scores and slightly higher fractions for higher infection probabilities. The analysis of the PTR shows that in general the efficiency of the testing for detecting infected asymptomatics is better for the strategy based on testing cohabitants only (ECS2) than for the ones based on random testing (ECS1) or testing traced contacts (ECS3). In any case, as it was previously mentioned, the effect of these testing campaigns has not a huge impact in the infection containment. The strategies in the latter group, ECS4 and ECS5, are able to effectively keep the MIR below 30-40% at the cost of a higher MQR.

**Figure 4:**
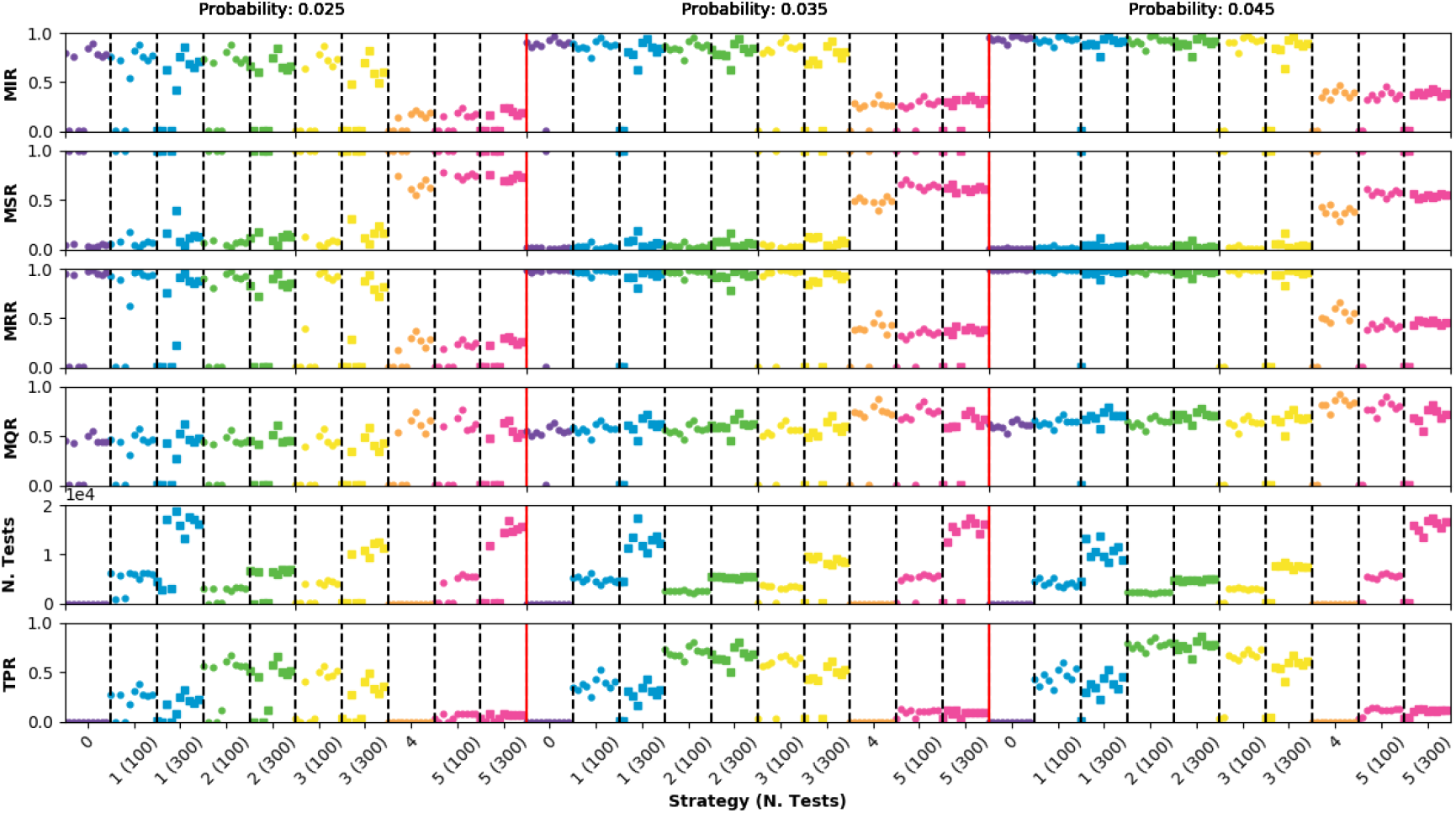
Epidemic observables for an infectious period of 4 days and a fraction of asymptomatic population of 25%. Points very close to 0 in the MIR observable indicate that the epidemic was controlled by the ECS.

### Dependency on the number of asymptomatic agents

Figure 5 shows the score of the different ECS for two scenarios with 25% and 75% of asymptomatic agents respectively. The infection probability has been fixed to 0.035 and the IPS to 4 days. A detailed inspection of these scenarios and the comparison among ECS reveal that there is a clear dependency on whether the first infected agents are asymptomatic since, contrary to the 25% case, in the 75% scenario the epidemic cannot be controlled with any strategy. The conclusions for the 75% scenario regarding the comparison among ECS is similar to what was discussed in the previous subsection, being the strategies based on massive quarantine the only ones making a real impact on the disease propagation. For all ECS, the MIR is higher in the 75% scenario. For ECS with massive confinement this rate increases about 20%. This was expected, since the agents not presenting symptoms will be freely infecting other people until they are tested or sent into quarantine.

**Figure 5:**
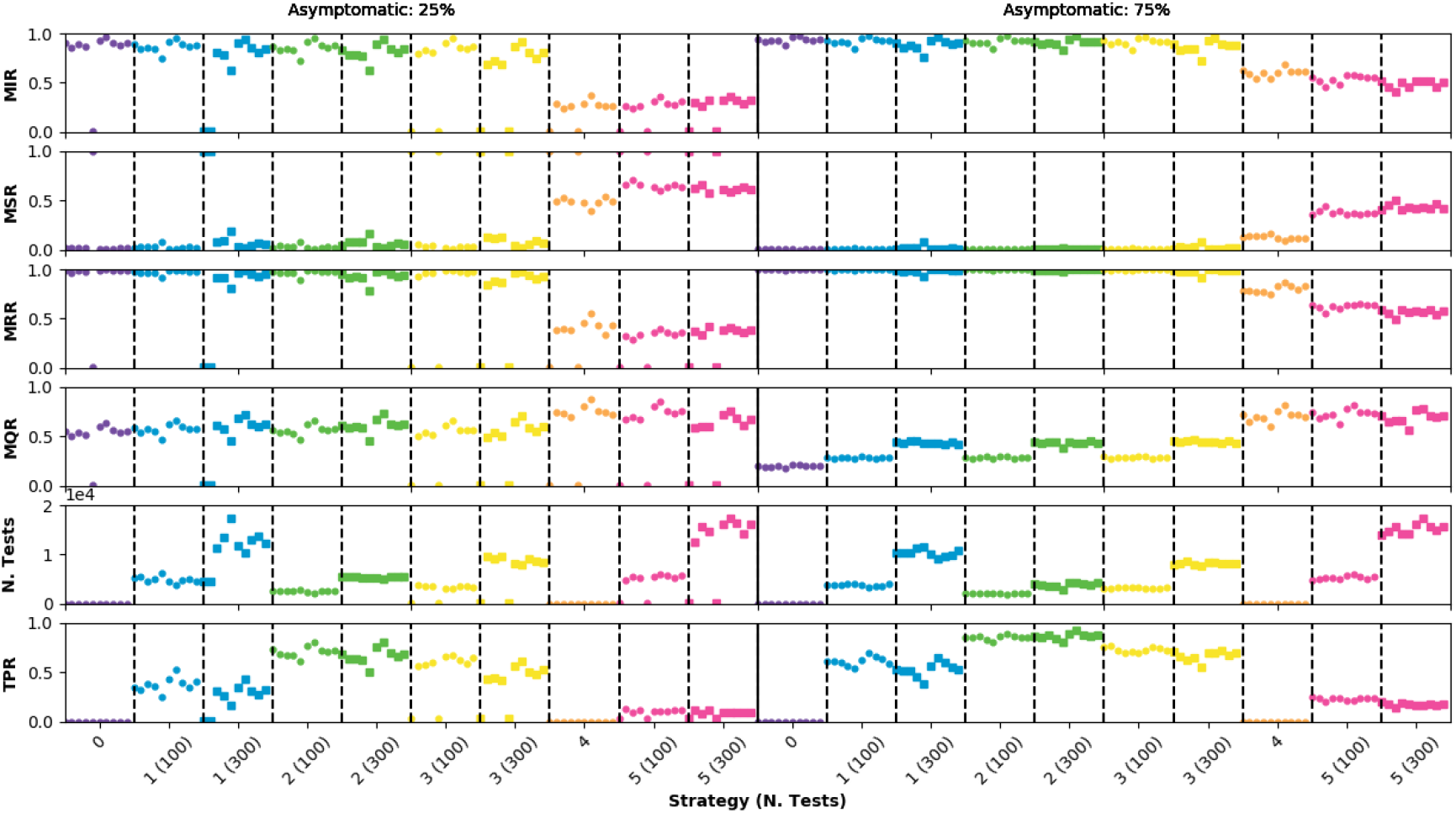
Epidemic observables for an infectious period of 4 days and an infection probability of 0.035. Points very close to 0 in the MIR observable indicate that the epidemic was controlled by the ECS.

### Dependency on the infectious period before symptoms appearance

Figure 6 shows the observables for different values of the IPS using an infection probability of 0.025 and 25% of asymptomatic agents. The dynamics of the epidemic are strongly affected by the IPS parameter. The case in which the IPS is zero, meaning that symptoms manifestation (if any) and infectiousness occur at the same time, results in epidemics that are always contained by any of the ECS. For an intermediate IPS of 2 days, the MIR scales to close to 50% for ECS0, with a trend inversely correlated with the ECS number. In the case of ECS3 with 300 hundred tests the epidemic can still be controlled in most of the cases. The ECS4 and ECS5 are also able to contain the epidemic, as expected, with a higher MQR. For an IPS of 4 days, the results analyzed in the previous sub-section are recovered. This study shows the capital importance of the IPS in the evolution of an epidemic and in the choice of ECS.

**Figure 6:**
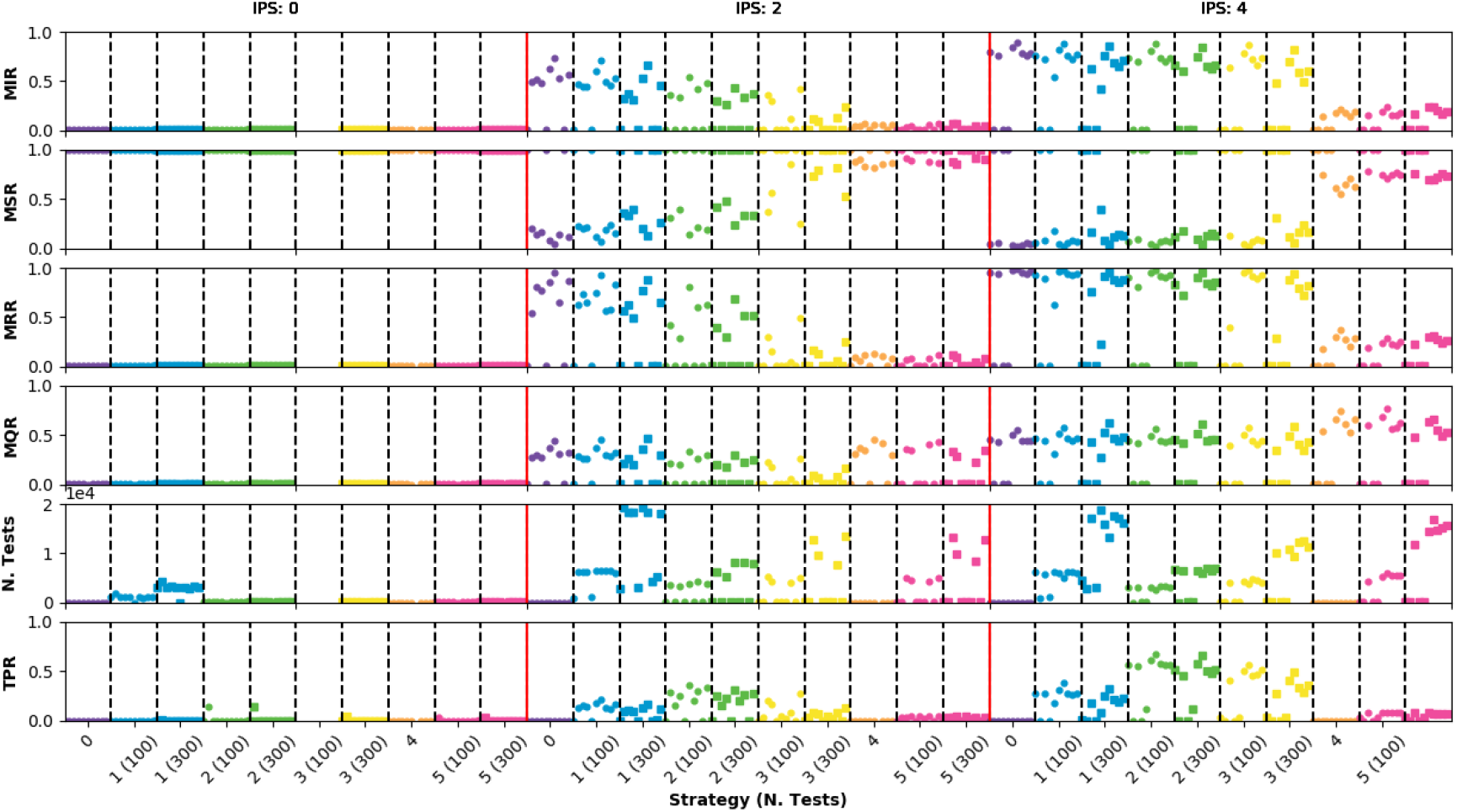
Epidemic observables for an infection probability of 0.025 and a fraction of asymptomatic agents of 75%. Points very close to 0 in the MIR observable indicate that the epidemic was controlled by the ECS.

### Dependency on the CTS engagement

Figure 7 shows the observables for different values of the population engagement to the CTS (100%, 75% and 25%). The infection probability has been fixed to 0.035 and the fraction of asymptomatic agents to 25%. No change is expected for ECS 0 to 2, since they are not considering CTS information. For the others, a clear loss of performance is observed in all cases with respect to the full engagement scenario. The ECS4 and ECS5 can achieve a containment at the level of 10-15% in the MIR observable with full engagement. However, these values degrade to the level of 80% in the case of an engagement of 25%, showing essentially no difference with the other ECS. In practice, this means that for low engagement scenarios using CTS information is not better than ECS doing tests on cohabitants or even randomly.

**Figure 7:**
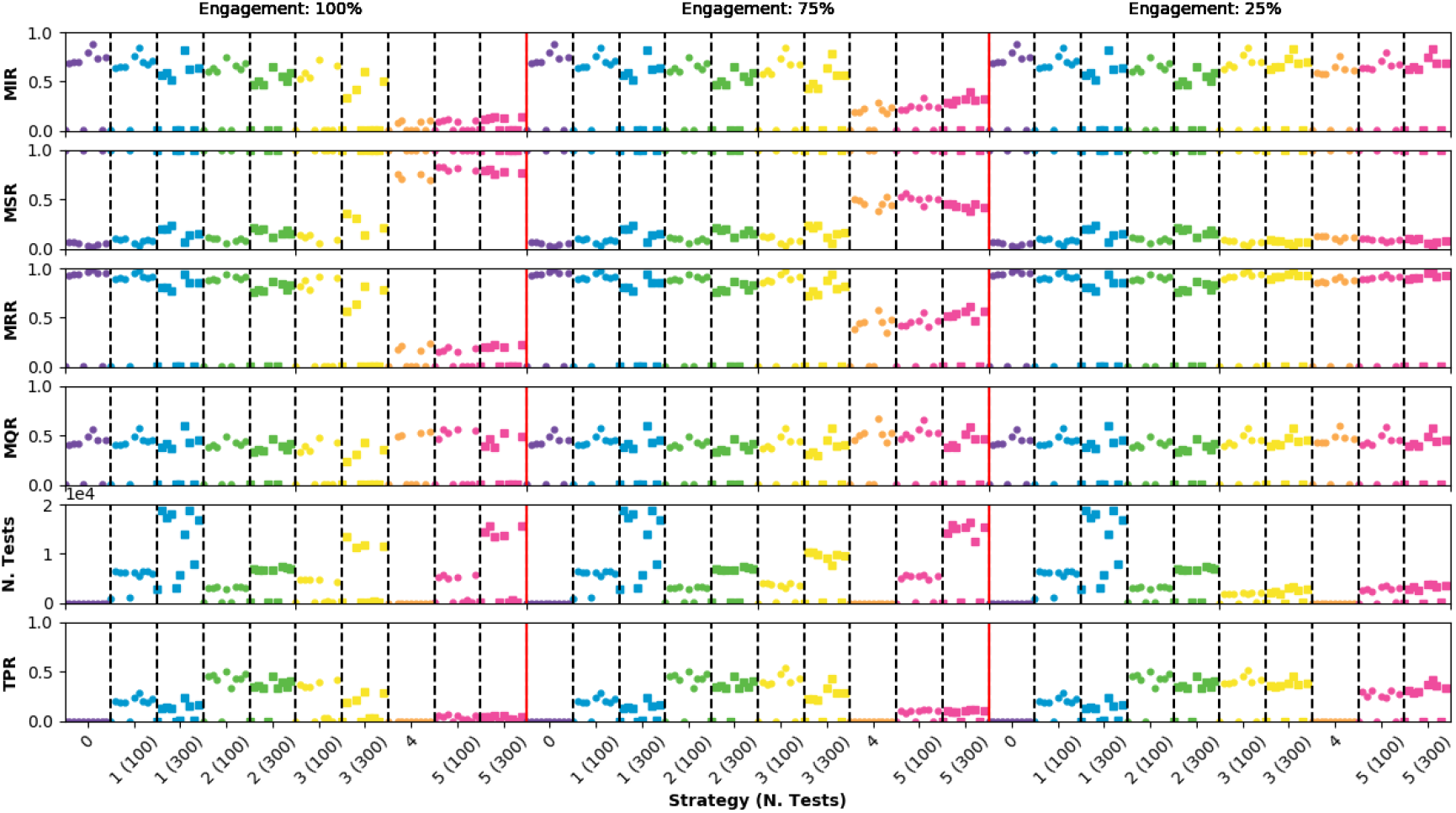
Epidemic observables for three CTS engagement values (100%, 75% and 25%). Infection probability has been fixed to 0.035 and the fraction of asymptomatic agents is 25%. Points very close to 0 in the MIR observable indicate that the epidemic was controlled by the ECS.

## 5 Parameter estimation using a graph database

The graph database has proven to be very effective in the automation of the ECS (regardless the success of the strategy itself). The data stored in the database allows to extract useful information to understand the progression and characteristics of the epidemic. For instance, if a CTS can log in the times and locations of the contacts to the database, probabilities of infection at different places and times can be estimated. Other properties of the epidemic can also be obtained by applying statistical methods to the data. In the following lines, we provide a mathematical framework to perform parameter estimation using the information of a graph database with CTS information.

An habitat as described before is considered in which the probability of an agent infecting another is constant in time whenever the two agents are closer than a predefined distance. A CTS is present recording any proximity contact between two agents. This exercises considers the best case scenario, in which a full adoption of this system among the agents is assumed, no asymptomatic agents are present, and the IPS is set to 0.

According to the model, the time is considered to be discrete and to evolve with a minute granularity. Under these assumptions, after a number T of minutes, the probability of an agent not being infected can be expressed as (1 − *p*)*^N^* where p is the probability of being infected in a minute when an infected agent is closer than the threshold, and where N is the total number of minutes in which the agent has been in this situation. If, at time T+1, a measurement is made on the system to see which agents are infected, the probability of being infected (y = 1) or not infected (y = 0) can be written as

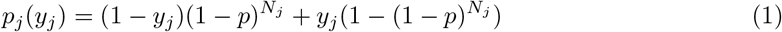

where N*_j_* is the number of minutes with another infected agent. This opens the possibility to define a likelihood function as follows:

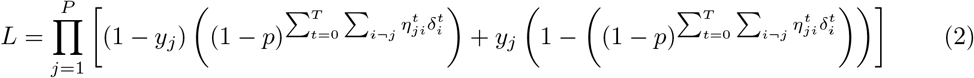

where 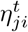 is 1 if there was a proximity contact between agents j and i at time t, and 0 otherwise, and where 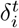 is 1 if agent i was infected at time t, and 0 otherwise. Taking into account that infections are continuous in time and determined by the infection and recovery times, the expression can be simplified by using a function Φ(*t*; *t_inf_*, *t_rec_*) that returns 1 if t is in between the two times and 0 otherwise. Then the expression reads:

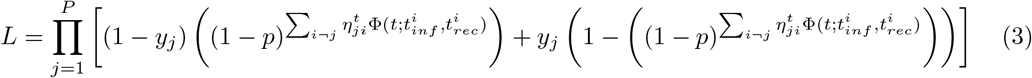

This quantity can be maximized (or alternatively q = -2 log(L) minimized) against unknowns in the system such as the infection and recovery times for a subset of agents and other properties. This procedure has been applied to estimate the infection probability in a small habitat of 1028 inhabitants. A per-minute probability of *p* = 0.0017 has been used in the model. Figure 8 shows the values of the q-statistic for different values of p.

**Figure 8:**
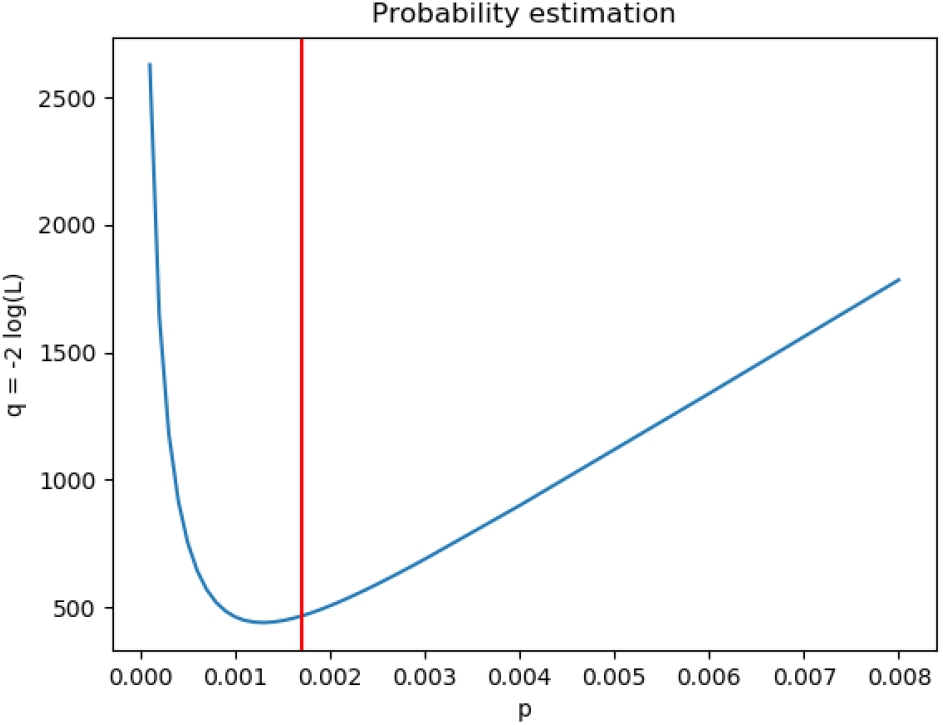
Value of the q-statistic associated to the likelihood as a function of the probability. The nominal value has been indicated with a red vertical line.

## 6 Conclusions

This work shows the strong dependency of the epidemic control strategies on the infectious period before the presence of symptoms and on the percentage of asymptomatic agents. This highlights the importance of properly estimating these parameters in order to make decisions on the planning and design of containment strategies. For large infectious periods before the manifestation of symptoms only the massive containment strategies (those implying a greater amount of people in quarantine) are capable of having a significant impact on the infection propagation. For low or intermediate infectious periods before the presence of symptoms and low or intermediate infection probabilities, the use of selective testing, either based on cohabitation or on traced contacts, can be useful. Under these scenarios, it has been observed that, even if testing families presents a higher positive testing rate, the strategies based on tracing contacts with high engagement are more effective since more test are performed. Additionally, it has been observed that a low engagement of the population to the contact tracing system largely degrades the performance of the strategies. In this scenario, random or cohabitant testing are as effective as contact tracing based strategies. According to the level of engagement (well below 25%) observed in countries where these tracing systems have already been used, the impact of these technologies on the epidemic control seems very limited.

Finally, this work offers a methodology to implement an epidemic observatory using a graph database. An example on how the stored data can be statistically treated to measure valuable properties of the epidemic has been shown. We recommend a further exploration and expansion of these methods in order to improve the response against an epidemic.

## Data Availability

All the data used in this work have been generated using Monte Carlo simulations.

## 7 Acknowledgements

We acknowledge the support of the Particle Physics group (CMS Tier-2 project with reference FPA2016-78727-R) and the Advanced Computing and e-Science group at the Institute of Physics of Cantabria (IFCA-CSIC-UC). We also acknowledge the Ramon y Cajal program of the Spanish Ministry of Science. Finally, we want to thank the company Muon Systems for their computational resources and Prof. Francisco Matorras and Arturo Medela Ceballos for their support and fruitful discussions.

## Notes

### Competing Interest Statement

The authors have declared no competing interest.

### Author Declarations

Institutional reviewed by IFCA

